# Navigator-gated free-breathing joint T_1_-T_2_ mapping of the kidney

**DOI:** 10.1101/2025.08.17.25333832

**Authors:** Pauline Calarnou, Augustin C. Ogier, Christopher W. Roy, Jean-Baptiste Ledoux, Angela Rocca, Menno Pruijm, Roger Hullin, Jean-Paul-Vallée, Jérôme Yerly, Ruud B. van Heeswijk

**Affiliations:** Department of Radiology, Lausanne University Hospital and University of Lausanne, Lausanne, Switzerland; CIBM Center for BioMedical Imaging, Lausanne and Geneva, Switzerland; Cardiology Service, Cardiovascular Department, Lausanne University Hospital and University of Lausanne, Lausanne, Switzerland; Nephrology Service, Internal Medicine Department, Lausanne University Hospital and University of Lausanne, Lausanne, Switzerland; Radiology Service, Department of Diagnostics, Geneva University Hospital and University of Geneva, Switzerland

## Abstract

**Purpose:** To develop and evaluate a free-breathing 2D radial joint T₁-T₂ mapping technique for the kidneys at 3T, and to assess the impact of navigator gating parameters on mapping precision and accuracy.

**Methods:** The PARMANav sequence (PArametric Radial MApping with Navigator gating) was implemented for renal imaging, using 25 single-shot radial gradient echo acquisitions with five repeated magnetization preparations and lung-liver navigator gating to avoid through-plane motion. Virtual compressed coil and compressed sensing with spatial and contrast regularization was used for image reconstruction, followed by a model-based registration. An acquisition-specific joint T₁-T₂ dictionary was generated using extended phase graph simulations. T_1_ and T_2_ accuracies were quantified in a phantom study versus gold standard spin-echo-based sequences. The influence of the navigator acceptance window width (NAWW) and navigator rejection on T_1_ and T_2_ precision were established in 10 healthy volunteers and were compared to routine T_1_ and T_2_ mapping. Three patients were scanned to demonstrate clinical feasibility.

**Results:** In the phantom, PARMANav T_1_ and T_2_ values showed high accuracy with the gold standard T_1_ and T_2_ values and were insensitive to rejected navigators (< 5% variation for T_1_ and T_2_). As expected from previous studies, in-vivo renal PARMANav T_1_ and T_2_ values were higher than routine values but showed lower variability, both per subject and between subjects: in the cortex PARMANav T_1_=1601±48ms/T_2_=90.8±5.0ms vs routine T_1_=1307±108ms/T_2_=73.3±8.0ms, while in the medulla PARMANav T_1_=2044±82ms/T_2_ =90.3±5.4ms and routine T_1_=1560±122ms/T_2_=67.6±5.8ms. No T_1_ or T_2_ trend was observed for the different NAWW. Feasibility was demonstrated in patients, where high-quality maps were obtained.

**Conclusion:** PARMANav allows for precise and accurate joint T_1_-T_2_ mapping of the kidneys without requiring breath holding. Through-plane motion artifacts were avoided with a navigator, which did not impact the accuracy or precision of the resulting maps.

## Introduction

In kidney disease, the T_1_ and T_2_ relaxation times reflect underlying changes in tissue composition,^1^ while a correlation between T_1_ cortico-medullary differentiation (CMD) and renal function has been demonstrated in several renal diseases.^2,3^ T_1_ CMD was proven to be related to the degree of fibrosis, while elevated cortex T_1_ values is strongly associated with poor renal outcome in patient with chronic kidney disease and with allograph kidneys,^4^ and oedema and T_2_ increases with inflammation and edema.^5^ Renal T_1_ mapping is typically performed with modified Look-Locker inversion recovery (MOLLI),^5,6^ which is known to underestimate the T_1_ value.^7^ Renal T_2_ mapping is less common and is usually performed with a turbo spin-echo sequence, which typically overestimates the T_2_ value.

Instead of mapping a single relaxation time per acquisition, over the past decade, multiparametric mapping techniques have enabled the simultaneous measurement of multiple relaxation times in a single acquisition. These approaches have gained interest thanks to their ability to provide more comprehensive insights into renal structure and function,^5^ while known pulse sequence imperfections (e.g., slice profile, inversion efficiency) can be incorporated in the fitting model to improve accuracy. Although such techniques have been successfully commercialized for neuroimaging with MR fingerprinting,^8^ and a large number of variations has been studied for cardiac MRI,^9–12^ its application to renal imaging is thus far limited to the research setting, partly because they require breath holding, which is not always feasible in kidney disease patients. The development of free-breathing techniques would represent an impactful step forward towards the integration of T_1_ and T_2_ mapping in clinical practice to assess kidney structure in these patients. Recent studies have demonstrated applications for breath-held joint T_1_-T_2_* mapping^13^,T_1_-T_2_ mapping^14^, and free-breathing T_1_-T_2_* mapping^15^ of the kidney at 3T.Most of the abovementioned techniques require breath holding, which is not always feasible in patients. Free-breathing archieved with respiratory gating,^15^ relies on the assumption of a constant respiratory cycle duration and used a two parameters analytical fit, that doesn’t allow to model precisely the magnetization evolution.

Conversely, many recent multiparametric techniques claim to allow for free breathing through the use of state-of-the-art in-plane retrospective motion correction (i.e., registration) between the source images, but do not account for through-plane motion: motion in the direction perpendicular to the image plane and thus invisible to a motion correction algorithm. This omission could be significant, potentially leading to perceived increased values in healthy tissues or normal values in a lesion. The reliance on registration, especially non-rigid registration, could also induce additional errors that propagate in final maps. To enable a free-breathing acquisition that avoids such discrepancies between its source images, a lung-liver navigator can be used to track the diaphragm position,^9^ albeit at the cost of acquisition efficiency. The accuracy of such a technique was previously assessed in a cardiac numerical phantom and reported no dependencies on the number of navigator rejections.^16^

In the current study, we aimed to demonstrate that an adaptation of the free-breathing navigator-gated multi parametric mapping technique PARMANav (PArametric Radial MApping with Navigator gating)^16^ can be used to obtain accurate and precise parametric maps of the kidney at 3T while avoiding through-plane motion.

## Methods

### Pulse sequence design

We adapted PARMANav^16^ for renal imaging by implementing a free-breathing acquisition of 25 magnetization-prepared single-shot 2D gradient-recalled echo (GRE) images, each acquired with a continuous golden angle trajectory (Figure 1). To enable magnetization recovery, the blocks (i.e., a magnetization preparation and a single-shot image acquisition) were repeated every 1s. To achieve joint T₁-T₂ sensitivity, five different contrast preparations were repeated in series: one adiabatic inversion pulse (a 5.12 ms hyperbolic tangent), no preparation, and three different T₂-preparation modules. This set was repeated five times to ensure contrast diversity throughout the different number of skipped navigators (Figure 1 A).

**Figure 1.**
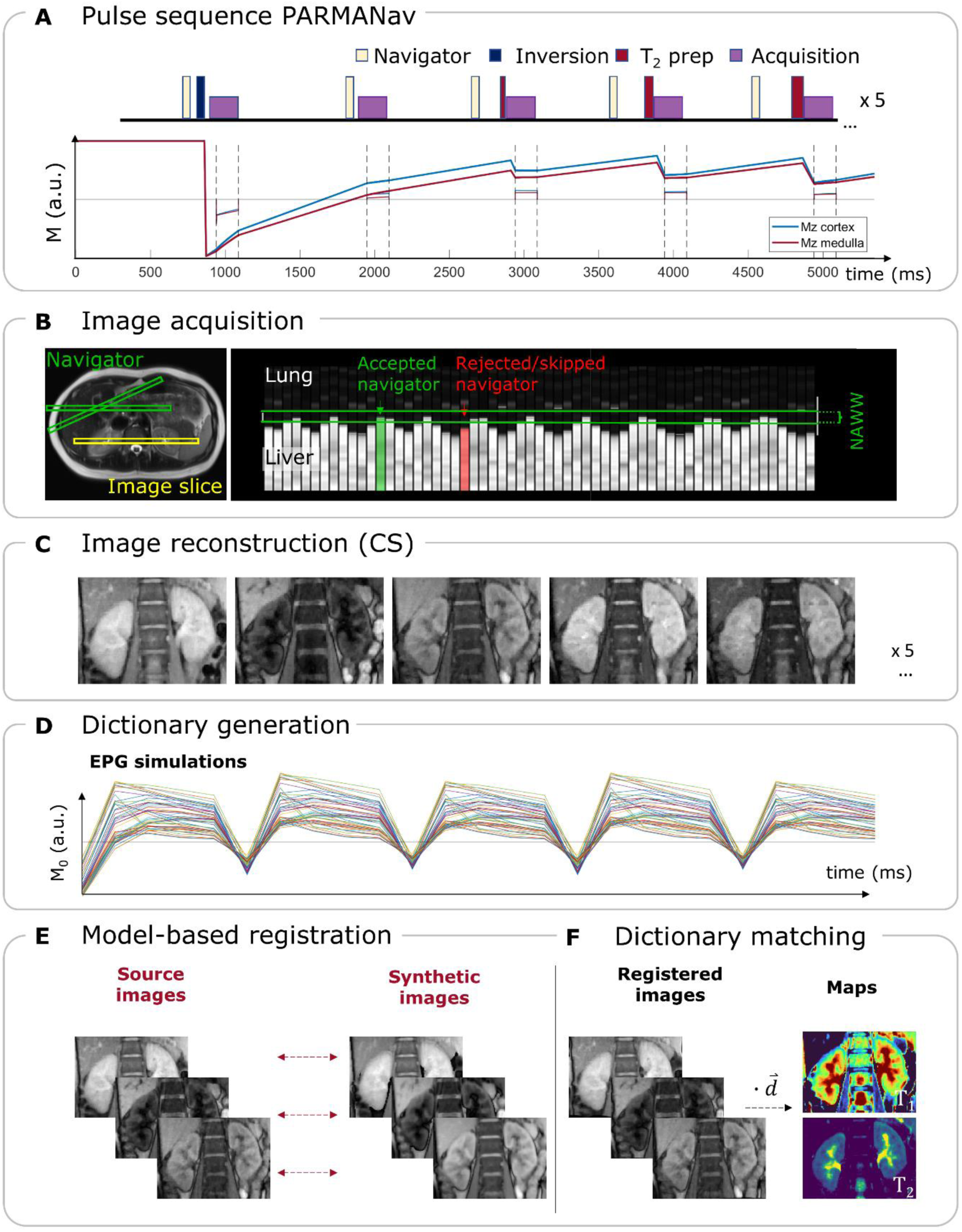
Overview of the free-breathing radial 2D joint T_1_-T_2_ mapping technique PARMANav. **A)** The pulse sequence diagram shows the first of five repeated blocks that consist of five differently prepared images, and the simulated magnetization of a healthy renal cortex and medulla. **B)** Illustration of the placement of the navigator (green) and image slice (yellow) as well as a trace of the navigator with the acceptance window as two parallel green lines. **C)** Example first 5 images in a healthy volunteer kidney, reconstructed using compressed sensing (CS). **D)** The dictionary is created through EPG simulations of the magnetization across the 25 images. **E)** Model-based motion correction between source and synthetic images obtained with the dictionary and the first unregistered maps. **F)** The final maps are obtained by computing the pixel-wise dot-product between the registered images and the dictionary 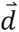.

Respiratory motion was tracked using a lung-liver navigator acquired before each preparation, with slice tracking enabled. To ensure that the timing of all magnetization changes was known, both preparation and readout modules were skipped when navigator rejection occurred.

All data were acquired on a 3T clinical scanner (Magnetom Prisma or PrismaFit, Siemens Healthineers, Forchheim, Germany) with nominal matrix size 192×192 (resulting in a 384×384 matrix through oversampled radial gridding), 45 continuous golden-angle (68°) radial lines per image (corresponding to 15% radial Nyquist sampling), pixel size=(1.56mm)^2^, slice thickness 8mm, flip angle 12°, bandwidth 789Hz/pixel, repetition time TR 3.49ms, echo time TE 1.56ms, acquisition window duration 151ms, inversion time TI 68ms (for the image directly after the inversion), and T_2_-preparartion modules echo times 23/45/70 ms. A fixed 5s delay at the start of the pulse sequence allows for complete magnetization relaxation between successive acquisitions. For all acquisitions we used a 34-channel chest-spine coil array.

### Image and map reconstruction

Individual RF coil elements were combined using region-optimized virtual (ROVir) coils^17,18^ to minimize radial streaking artifacts coming from the arms and the abdominal fat. Briefly, the 192×192-pixels central part of the acquired image was automatically selected as the region of interest, and the periphery of the image was designated as the unwanted signal region. A generalized eigenvalue decomposition was used to identify virtual coils that maximize the signal-to-interference ratio (SIR). The smallest set of virtual coils capturing ≥90% of the total signal energy was retained.

From these undersampled k-spaces, images were reconstructed using compressed sensing with total variation regularization in the spatial dimension and local-low-rank regularization along the contrast dimension:^19^

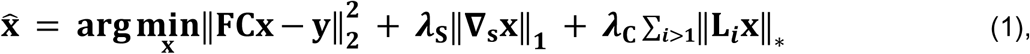

where **X̂** is the reconstructed image, **F** is the nonuniform fast Fourier operator, **C** is the coil sensitivity, **y** refers to the acquired k-space, **∇**_**s**_ is the first-order difference operator along the spatial dimension, **L**_***i***_ is the operator that extracts the *i*-th spatial patch of size 6×6×6 pixels from **x** and forms a Casorati matrix with the contrast dimension, ‖·‖_∗_ is the nuclear norm, and *λ*_*S*_ and *λ*_*C*_ are the corresponding regularization weights along the spatial and contrast dimensions, respectively. Regularization parameters, *λ*_*S*_ = 0.01 and *λ*_*C*_ = 0.06 were empirically selected for optimal trade-off between undersampling artifact removal and image blurring.

Considering individual navigator rejections, an acquisition-specific signal dictionary was generated via extended phase graph simulations in MATLAB (version R2023b, The Mathworks, Natick, Massachusetts, USA) across a wide range of T₁ values from 0 ms to 5000 ms in 10 ms increments, and from 5020 ms to 6000 ms in 20 ms increments. and T₂ values from 0 ms to 450 ms with a 5 ms increment and from 460 ms to 700 ms with a 10 ms increment, incorporating slice profile effects (discretized in 50 isochromats) and inversion inefficiency correction.^20^ The precise acquisition timing was extracted from raw data to reflect navigator acceptance. The magnetization was simulated at the center of each echo, and the signal averaged over each radial single-shot echo train, resulting in a 25×60 396 (T_2_>T_1_ cases being excluded) matrix of simulated complex signals that formed the dictionary.

A previously described model-based non-rigid image registration^16^ was applied to account for residual in-plane motion that resulted from residual differences in the respiratory phase while accounting for the strong contrast variations in these images (Figure 1 E). Here, a first set off maps was generated via pixel-wise dictionary matching without motion correction. Synthetic images with matched contrast and averaged motion were then created and used as references for non-rigid optical-flow-based registration.^21^ Final maps were obtained by repeating dictionary matching on the motion-corrected images.

### Phantom study

The “ISMRM/NIST” phantom^22^ (Premium System 130, CaliberMRI, Boulder, USA) was scanned to evaluate the accuracy of PARMANav readout echo trains were separated by intervals of 1s, which were sporadically extended to 2s or 3s such that navigator rejections could be emulated. Clinical routine T_1_ and T_2_ maps (pixel size=(1.4-1.9mm)^2^, slice thickness 8mm) were acquired using 5(3)5 MOLLI,^6^ and T_2_-prepared (T_2_-prep) bSSFP T_2_ mapping,^23,24^ respectively. It was compared against the values obtained with gold-standard inversion-recovery spin-echo (for T_1_ relaxation) and spin-echo (for T_2_ relaxation) techniques. The different sequences parameters are reported in Table 1.

**Table 1.**
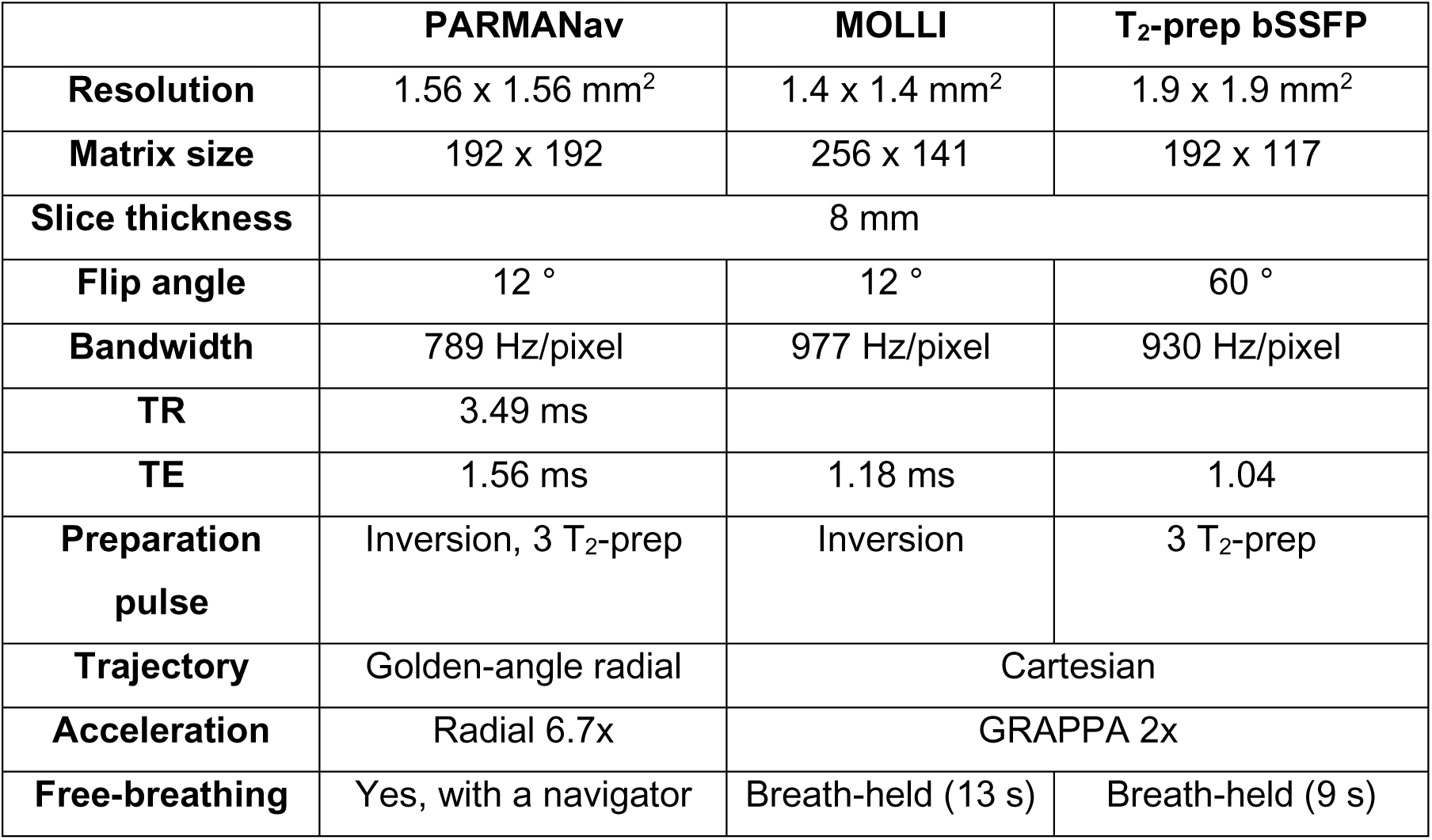
Parameters of the sequences used for T_1_ and T_2_ mapping. The same parameters were used in the phantom and in-vivo scans.

|  | **PARMANav** | **MOLLI** | **T<sub>2</sub>-prep bSSFP** |
| --- | --- | --- | --- |
| **Resolution** | 1.56 x 1.56 mm<sup>2</sup> | 1.4 x 1.4 mm<sup>2</sup> | 1.9 x 1.9 mm<sup>2</sup> |
| **Matrix size** | 192 x 192 | 256 x 141 | 192 x 117 |
| **Slice thickness** | 8 mm |  |  |
| **Flip angle** | 12 ° | 12 ° | 60 ° |
| **Bandwidth** | 789 Hz/pixel | 977 Hz/pixel | 930 Hz/pixel |
| **TR** | 3.49 ms |  |  |
| **TE** | 1.56 ms | 1.18 ms | 1.04 |
| **Preparation pulse** | Inversion, 3 T<sub>2</sub>-prep | Inversion | 3 T<sub>2</sub>-prep |
| **Trajectory** | Golden-angle radial | Cartesian |  |
| **Acceleration** | Radial 6.7x | GRAPPA 2x |  |
| **Free-breathing** | Yes, with a navigator | Breath-held (13 s) | Breath-held (9 s) |

### Healthy volunteer study

Ethics approval was obtained from the Ethics Committee of the Canton of Vaud (CER-VD) of Switzerland under reference numbers 2021-00697 and 2022-00934. All participants provided written informed consent to participate prior to enrollment.

PARMANav maps of N=10 healthy volunteers (31.8±9.1 y, 4F) were acquired with four different navigator acceptance window widths (NAWWs of ±4mm, ±8mm, ±16mmm, and ±32mm) of the lung-liver navigator in a randomized order to study the impact of through-plane motion. Volunteers were instructed to breath normally. Like in the phantom study, clinical routine breath-held T_1_ and T_2_ maps were acquired using MOLLI and T_2_-prepared bSSFP T_2_ mapping, respectively.

The T_1_ and T_2_ value of the visible cortex and medulla as well as the CMD ratio (as the ratio T_1cortex_/T_1medulla_) were determined for each map in each volunteer by manually segmenting regions of interest (ROIs) in both kidneys on the T_1_ map. Values from the two kidneys were averaged. The coefficient of variation (CoV) was calculated as the regional standard deviation divided by the mean relaxation time. The acquisition time was recorded for the four NAWWs.

### Patient study

To preliminarily evaluate its clinical feasibility, PARMANav was acquired in three patients (65.7±15.0,1F) as part of an ongoing study on heart failure.^25^ Two patients had chronic kidney disease (CKD), one with heart failure with preserved ejection fraction (HFpEF) and one with heart failure with reduced ejection fraction, while one patient had HFpEF but no CKD. In each patient, one map was acquired with a NAWW of ±8mm. No routine mapping techniques were acquired due to the constraints of the total MRI protocol duration. Anatomical reference images (T_2_-weighted HASTE) were acquired at the same location for two of the three patients, and at a slightly different orientation for the last patient.

### Statistics

Agreement between the measured and gold-standard values in the phantom was evaluated using the slope and coefficient of determination (R²) from linear regression analysis for both PARMANav with and without skipped navigator, and with the routine techniques. Bland-Altman analysis was performed to quantify biases and limits of agreement, and a paired t-test was conducted to evaluate statistical significance.

In the healthy volunteers, the T_1_ and T_2_ values of the cortex and medulla were extracted for the four NAWWs and the two routines methods. A Shapiro-Wilk test was performed to assess normality. Their differences, as well as the time of acquisition differences, were tested with a repeated-measures analysis of variance (RM-ANOVA) with post-hoc Tukey analysis. The regional T_1_-T_2_ values and CoV means and standard deviations (SDs) across the healthy volunteers were reported. Bland-Altman analysis was performed to quantify biases and limits of agreement.

## Results

### Phantom study

In the phantom, PARMANav showed high agreement with the gold standard, both with and without rejected navigators (Nskipped =23) (Figure 2): the slope of the correlation was closer to identity than the routine technique for T_1_ (1.05 and 1.00 for without and with skipped heartbeat, respectively vs 0.76 for MOLLI), while there was a larger difference for T_2_ (1.10 and 1.21 vs 0.60). R² was above 0.99 for both the PARMANav scans and MOLLI, and slightly different for the T_2_ mapping methods (>0.99 for the two PARMANav vs 0.94 for T_2_-prep bSSFP). As with previous studies,^16^ PARMANav with and without skipped navigators did not significantly differ for T_2_ (P>0.1). A significant difference was reported for T_1_ (P=0.005), with a small average relative difference (6.6%).

**Figure 2.**
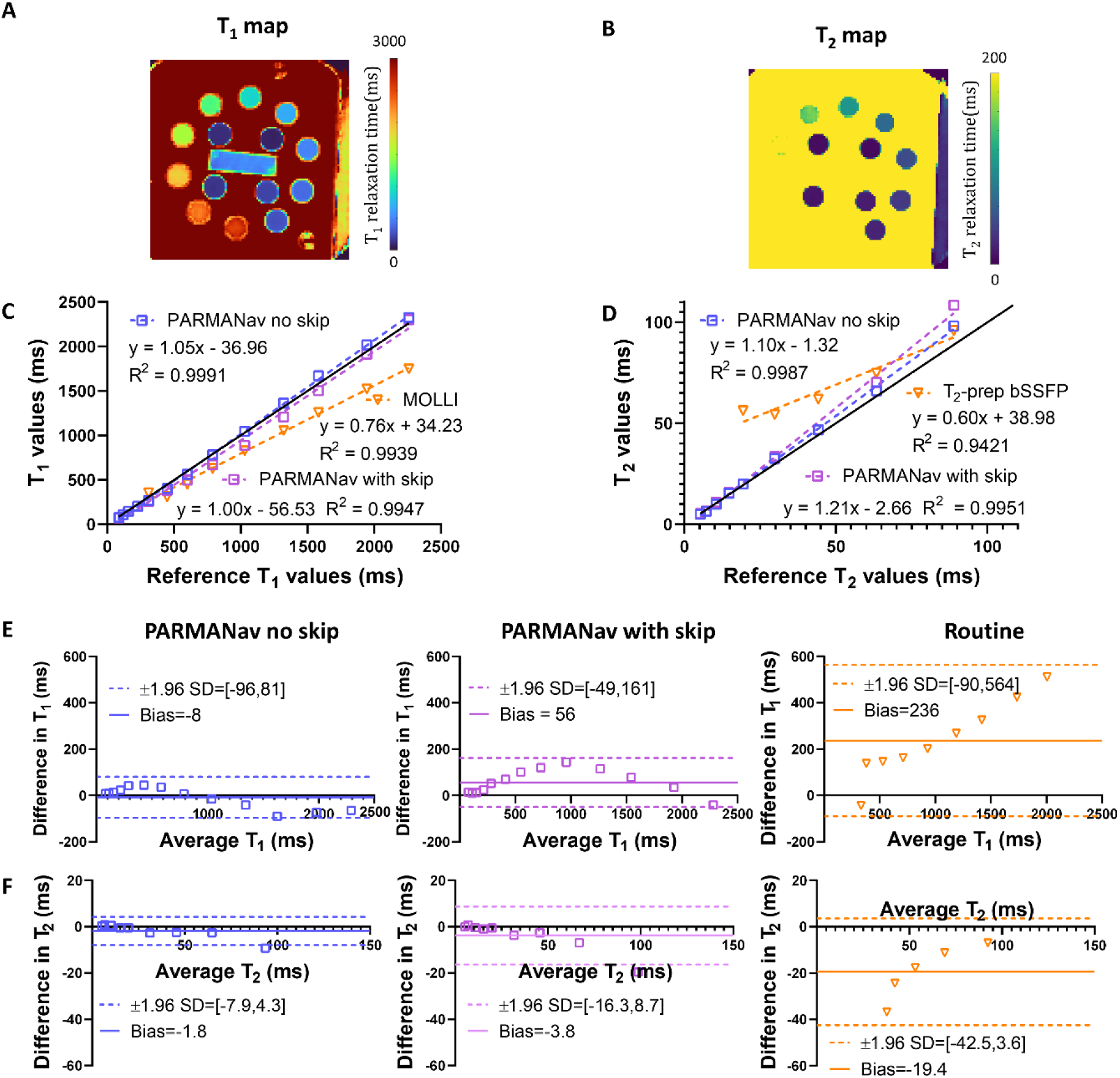
Agreement of PARMANav T_1_ and T_2_ maps in the ISMRM-NIST phantom compared to spin-echo reference values in case of 23 skipped navigator. **A)** T_1_ map of the phantom obtained with PARMANav. **B)** T_2_ map of the reference phantom obtained with PARMANav. **C)** Linear regression of PARMANav T_1_ values with and without rejected navigator and the clinical routine 5(3)5 MOLLI versus gold-standard IR-SE. **D)** Linear regressions of PARMANav T_2_ values with and without rejected navigator and the clinical routine T_2_-prep bSSFP in the phantom in the clinically relevant range versus gold-standard SE. **E)** T_1_ Bland-Altman plot of PARMANav with and without rejected navigator and MOLLI versus gold-standard IR-SE. The bias and confidence bounds are reported in the legend. PARMANav presented a larger bias when adding a large number of skips (N=23). **F)** Same as E) for T_2_ with T_2_-prep bSSFP as the clinical routine and SE as the gold-standard.

### Precision in Healthy Volunteers

In all 10 healthy volunteers, visually sharp and precise maps of the kidney were obtained with PARMANav (Figure 3). PARMANav T_1_-T_2_ values and CMD were significantly different from routine techniques, which are known to underestimate T_1_ and overestimate T_2_,^26^ but there were no significant differences as a function of the different NAWWs (Figure 4). The cortex and medulla T_1_ values presented a larger spread for NAWW=±4 mm than for NAWW=±8 mm with more outliers (SD_4mm_ = 117 ms and SD_8mm_ = 48 ms for the cortex). NAWW=±4 mm also resulted in a significantly longer acquisition time (p<0.02 for NAWW=±4 mm versus all the other NAWWs)(Figure 5). Given these results and the need for balance between low navigator acceptance (at small NAWW) and tolerated through-plane motion (at large NAWWs), NAWW=±8 mm mapping was chosen as best compromise for further analysis.

**Figure 3.**
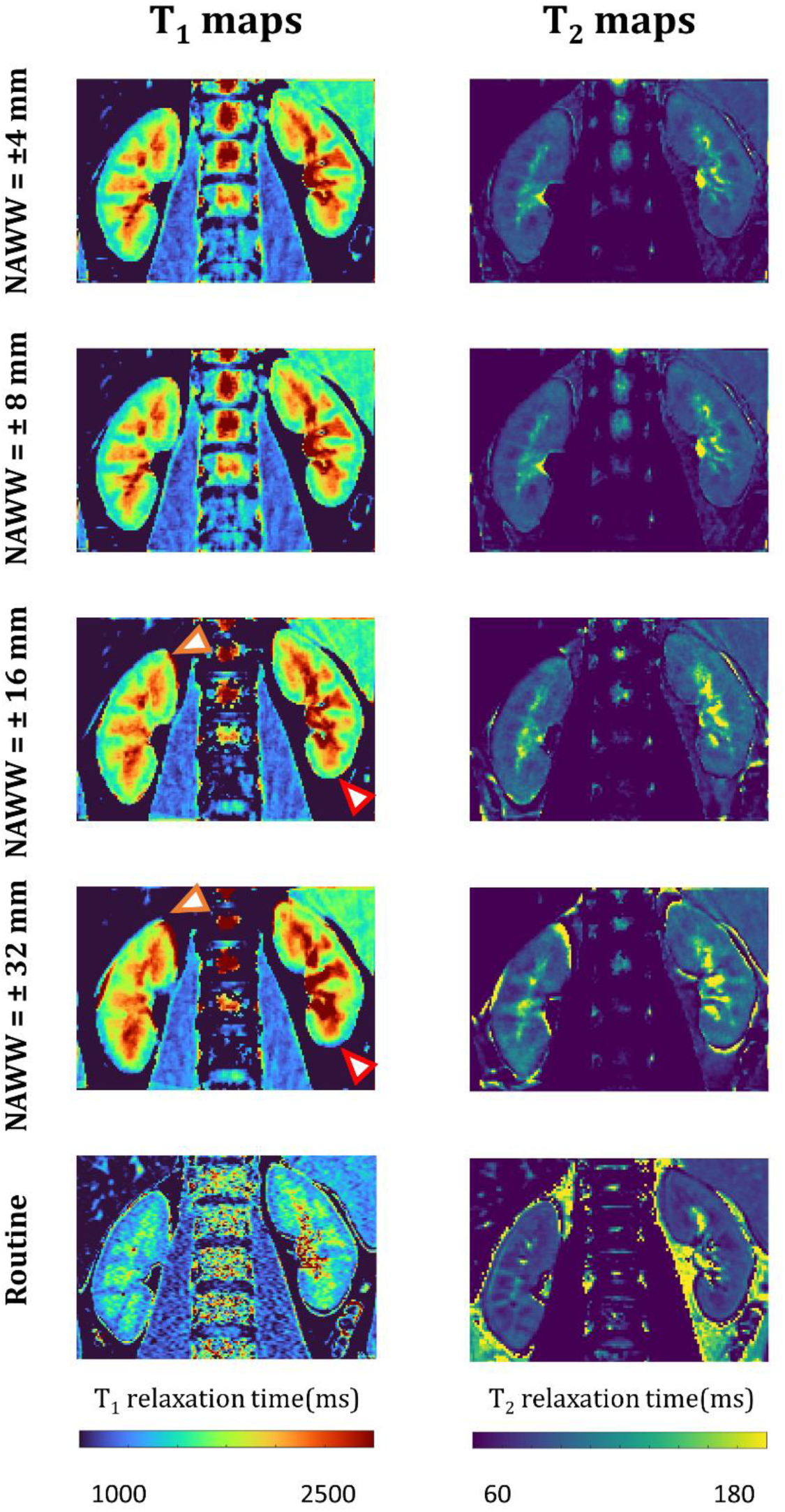
PARMANav T1 and T2 maps for the different NAWWs compared to the routine techniques. The NAWW did not significantly change the relaxation times in the resulting PARMANav T1 or T2 maps of the cortex, with the exception of the poles (red arrowheads), where partial volume effect lowered the T1 relaxation time. However, the shape and the T1 values of the medulla varied with the NAWW (orange arrowhead), likely due to through-plane motion. The PARMANav T1 and T2 values were consistently higher than those obtained with the routine techniques.

**Figure 4.**
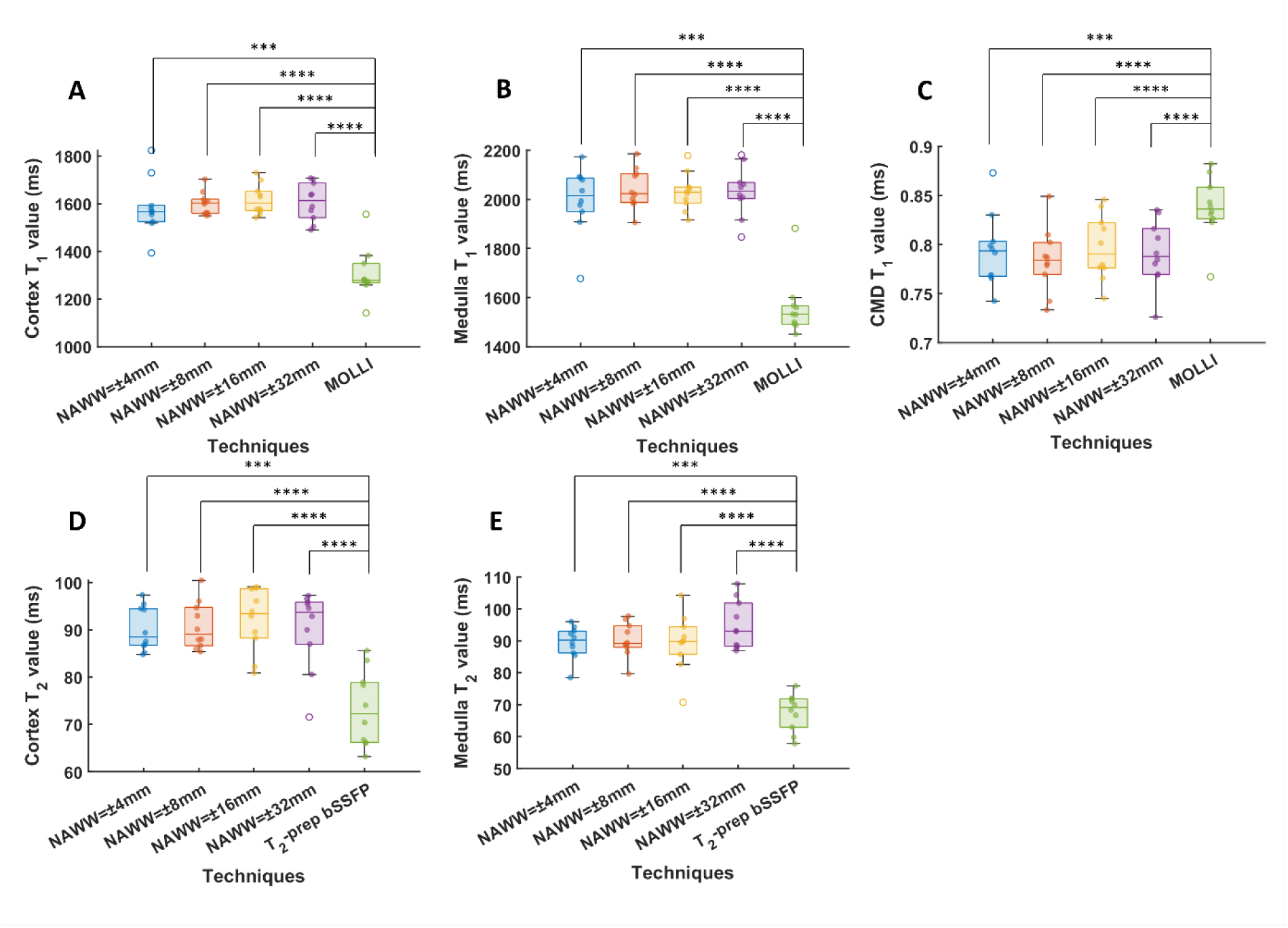
Impact of the navigator acceptance window width (NAWW). in 10 healthy volunteers. **A-C)** T_1_ values the cortex and in the medulla and the corresponding CMD ratio for the four NAWWs compared to MOLLI, which is known to underestimate T_1_ value, especially at these higher values. **D, E)** T_2_ values in the cortex and in the medulla for the four NAWW compared to T_2_-prepared bSSFP.

**Figure 5.**
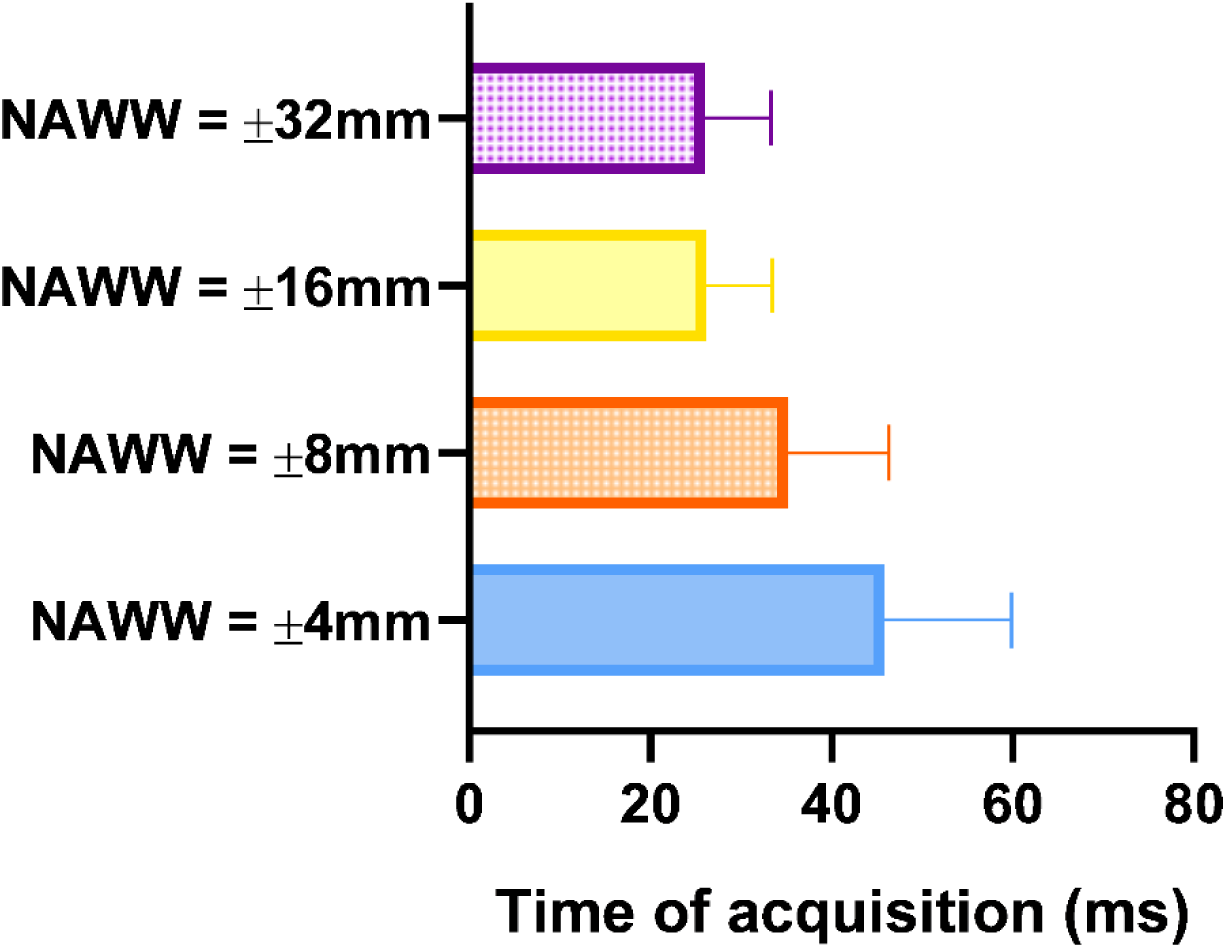
PARMANav acquisition time for the different NAWWs. Acquisition time was longer for smaller NAWW as expected. For NAWW=±16 and 32 mm, the time of acquisition was almost identical, due to no rejection for both NAWWs. The difference was significant between all the time of acquisition (p<0.02) except between NAWW=±16 and 32 mm (p=0.79)

PARMANav T_1_-T_2_ values were higher than routine techniques (Table 2). Smaller inter-subject SD was reported for all relaxation times.

**Table 2.**
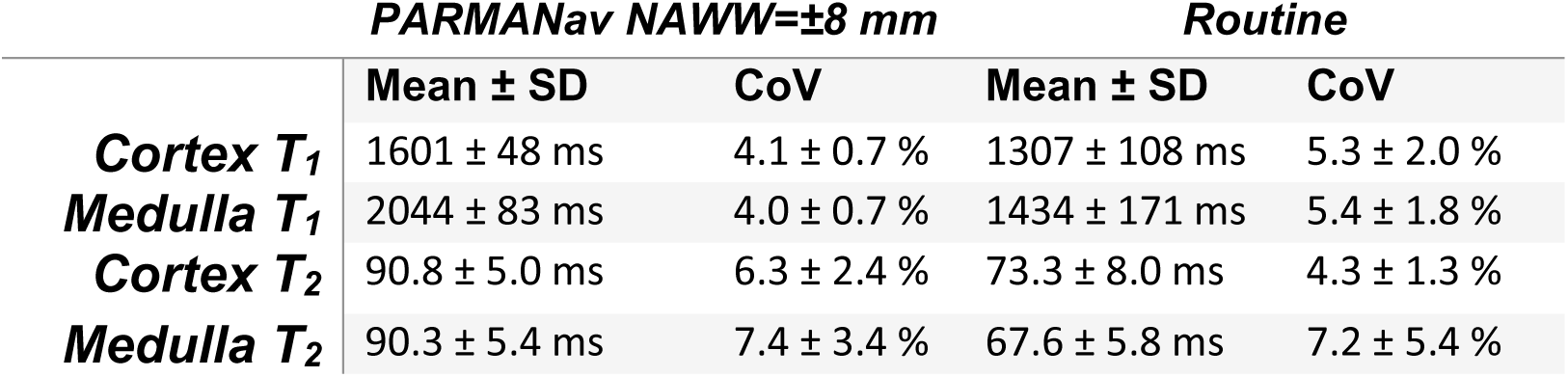
Cortex and medulla T_1_ and T_2_ mean and standard deviation in ms across the 10 healthy volunteers, as well as the coefficient of variation (CoV).

|  | PARMA Nav NAWW=±8 mm |  | Routine |  |
| --- | --- | --- | --- | --- |
|  | Mean ± SD | CoV | Mean ± SD | CoV |
| **Cortex $T_1$** | 1601 ± 48 ms | 4.1 ± 0.7 % | 1307 ± 108 ms | 5.3 ± 2.0 % |
| **Medulla $T_1$** | 2044 ± 83 ms | 4.0 ± 0.7 % | 1434 ± 171 ms | 5.4 ± 1.8 % |
| **Cortex $T_2$** | 90.8 ± 5.0 ms | 6.3 ± 2.4 % | 73.3 ± 8.0 ms | 4.3 ± 1.3 % |
| **Medulla $T_2$** | 90.3 ± 5.4 ms | 7.4 ± 3.4 % | 67.6 ± 5.8 ms | 7.2 ± 5.4 % |

The Bland Altman analysis (Figure 6) demonstrated a bias against the reference techniques, which is consistent with the phantom results and segmental averages.

**Figure 6.**
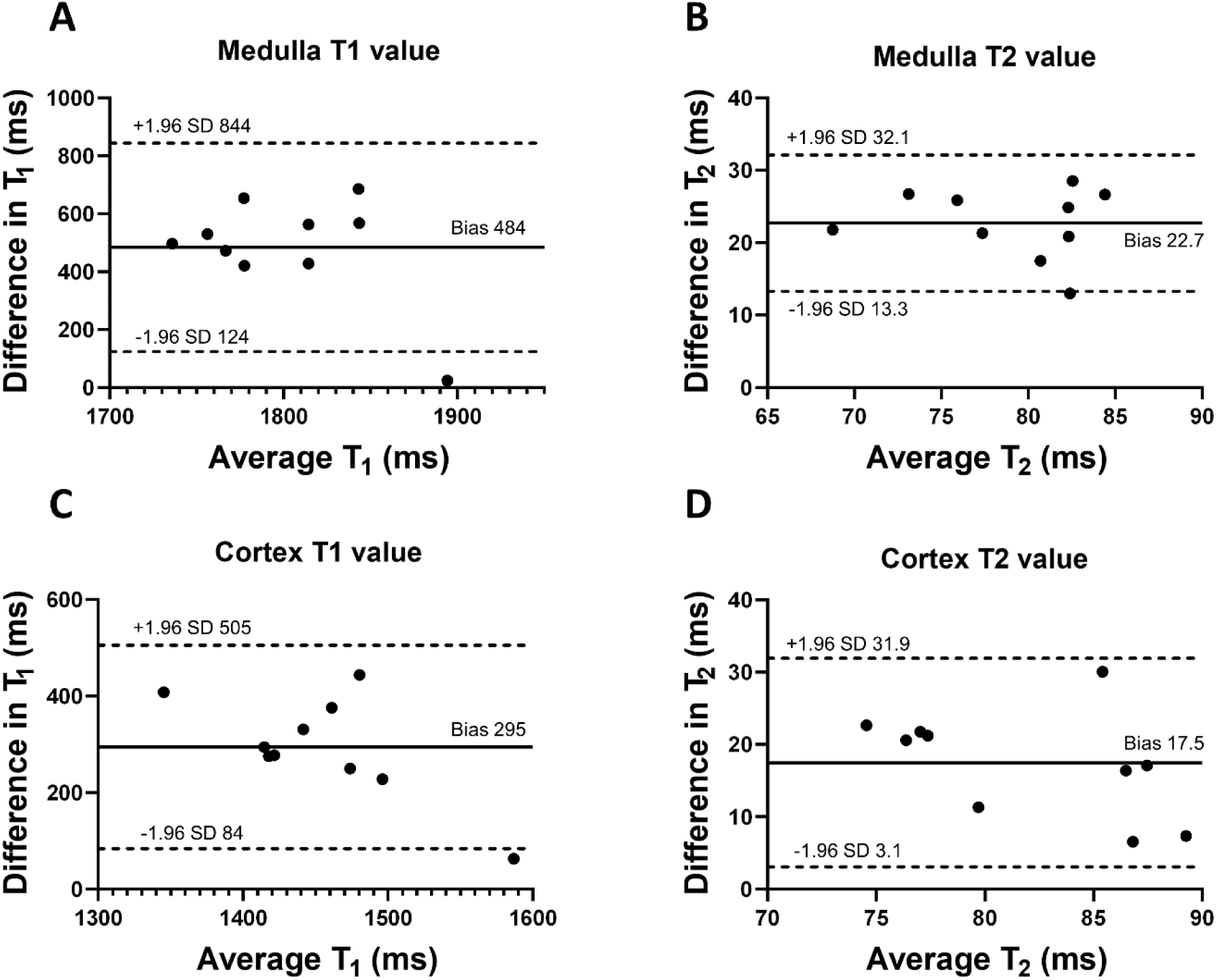
Bland-Altman analyses of the agreement between PARMANav with NAWW=±8mm and the reference techniques. **A)** Cortex T_1_ values. **B)** Cortex T_2_ values. **C)** Medulla T_1_ values. **D)** Medulla T_2_ values.

### Feasibility in Patients

PARMANav was successfully acquired in the three patients and resulted in sharp and artefact-free maps (Figure 7). A cyst, also visible on the localizer image could be clearly observed on the maps for Patient 1 – the free liquid interior results in very high T_1_ and T_2_ relaxation times. The individual structures in the kidney are more difficult to differentiate in all three patients, while they had a CMD ratio close to one (CMD ratio = 0.80,0.87 and 0.89 for patient 1,2 and 3, respectively) as expected.^2,3^

**Figure 7.**
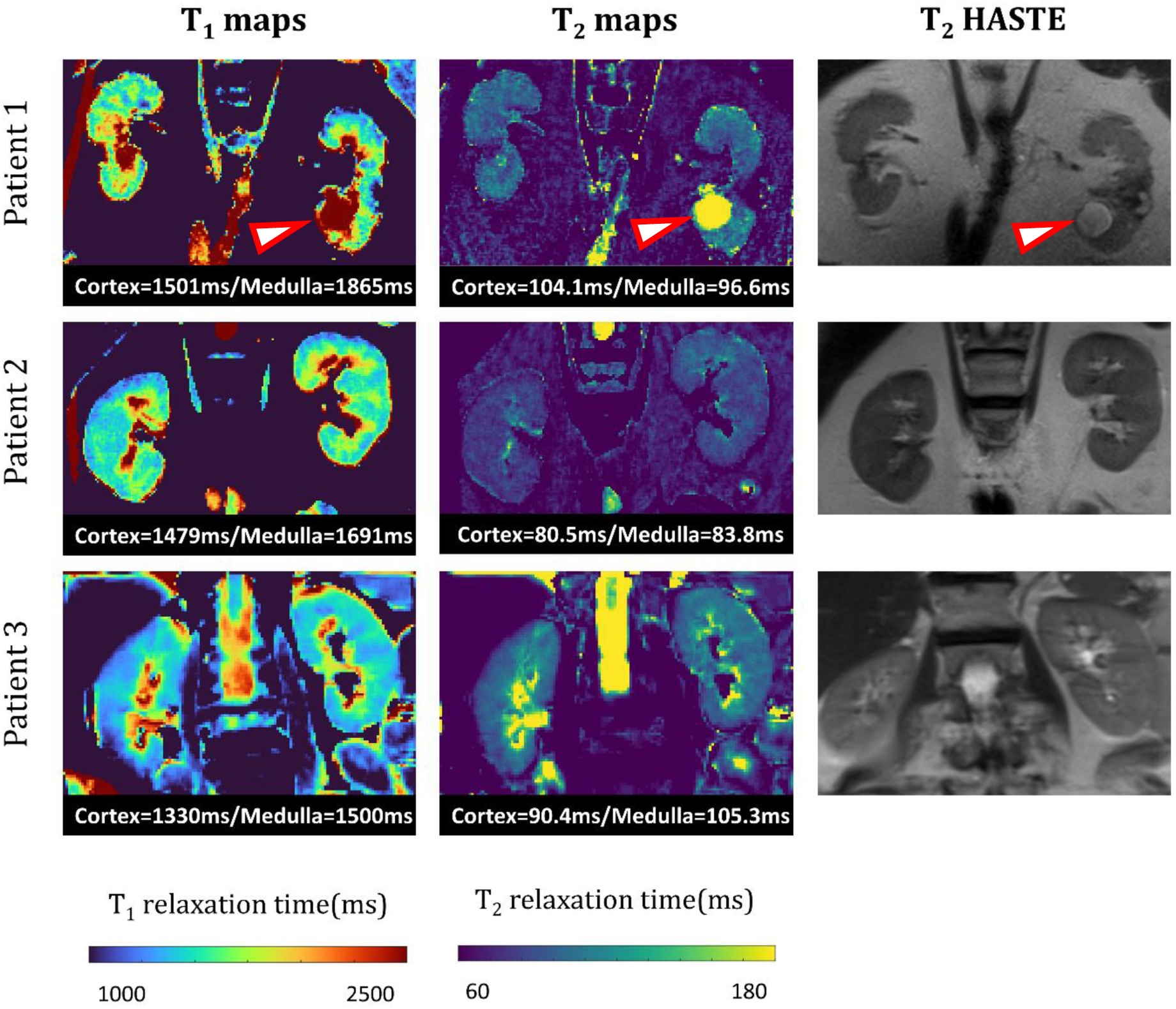
Parametric PARMANav maps obtained in patients. Patient 1 had chronic kidney disease (CKD) and heart failure with preserved ejection fraction (HFpEF) as well as a cyst (red arrowhead), Patients 2 had chronic kidney disease and heart failure with reduced ejection fraction, and Patient 3 had HFpEF without CKD. A T_2_ HASTE anatomical image is provided for anatomical reference.

## Discussion

In this work, a free-breathing 2D technique was characterized for joint T_1_–T_2_ mapping of the kidneys at 3T. Previous numerical simulations have shown that the estimated T_1_ and T_2_ values were not impacted by the number of rejected navigators^16^. The mapping phantom accuracy obtained in this study further support this finding.

PARMANav resulted in precise free-breathing joint T_1_-T_2_ maps of the kidney in the healthy volunteers. It was successfully acquired in all healthy volunteers with all tested NAWWs, which resulted in similar mapped relaxation times. In patients, free-breathing joint T_1_-T_2_ maps resulted in artefact-free maps of high quality with the added advantage that the T1 and T2 maps are intrinsically registered.

The Bland-Altman study in the healthy volunteers showed a bias compared to routine techniques, which was relatively high for the medulla T_1_ values, but agreed well with the phantom study. T_1_ and T_2_ values were systematically higher for PARMANav, both in the cortex and medulla, with large limit of agreement.

The higher T_1_ SD for NAWW=± 4 mm might indicate that a higher number of navigator rejections impacts the T_1_ precision. To mitigate this effect, a variable flip angle could be introduced for the acquisition, which could also be used to calculate a B_1_ map.^11^ Although the impact of the number and timing of rejected navigators (e.g., immediately following the inversion pulse) assessed in a mapping phantom would be valuable, the current experimental setup did not permit such precise control. Future studies using a motion phantom may enable these investigations and facilitate evaluation of accuracy in the presence of motion.

No consistent relationship was observed between NAWW and T_1_-T_2_ precision or accuracy, although this is likely due to avoiding clear artefactual borders during the segmentation process (as illustrated in Figure 3). This suggests that NAWW selection may be guided primarily by the trade-off between scan duration and through-plane motion tolerance. However, the shape of the cortex and medulla varied with the larger NAWWs, likely due to partial volume effects or limitations in the registration algorithm under conditions of increased motion. The increased number of outliers observed with NAWW=±4mm motivated the selection of an ±8 mm NAWW for subsequent experiments. Additional parameters, such as the flip angle and the number of radial lines per source image could be experimentally optimized and potentially reduced in future studies.

Several breath-held magnetic resonance fingerprinting (MRF) techniques have been proposed for renal mapping, all relying on Bloch equation simulations. Chen et al.⁸ developed an abdominal fingerprinting technique for simultaneous T₁ and T₂ mapping with B₁ correction. Compared to this method, which used spiral sampling and Bloch-based dictionary simulation, PARMANav yielded higher T₁ and T₂ values in both the cortex and medulla. More recently, Hermann et al.⁹ introduced a breath-held MRF sequence for T₁ and T₂ mapping across four slices. Their reported T₁ values, while still slightly lower, were more closely aligned with those obtained using PARMANav (T_1_ = 1456 ms and 1921 ms in cortex and medulla, respectively, vs. 1601 ms and 2044 ms with PARMANav). MacAskill et al.¹⁰ also presented a breath-held kidney MRF technique for T₁ and T₂ mapping with B₁ correction, again reporting shorter relaxation times than those measured using PARMANav. A free-breathing method was proposed by Ding et al.^15^ for T_1_ and T_2_* mapping, using respiratory triggering via a respiratory belt. The mapping was based on an analytical equation, which does not allow to model the imperfection of the acquisition (e.g., RF profile, inversion efficiency) and assumes a constant respiratory cycle duration.

The images acquired in patients were sharp and artefact-free. These results are encouraging and suggest that combined T_1_-T_2_ free breathing mapping with NAWW is possible in CKD patients, whether or not will they suffer from associated heart failure. As a next step, PARMANav should be compared to standard techniques. Future studies should also include a larger number of CKD patients, with different associated comorbidities known to lead to disturbances in breathing such as underlying lung disease or morbid obesity.

This study has several limitations. As mentioned, the number of included patients was small. Besides, dictionary-based multiparametric mapping is limited by discretization and computational demands: coarse grids introduce small errors, while finer grids increase generation and matching time. Other limitations of the proposed technique include challenges in sequence optimizations, as navigator rejections may have a greater impact on contrast variability than the parameter being optimized. Due to the complexity of kidney anatomy, only relatively small ROIs can typically be manually segmented. Although a second ROI was drawn on the second kidney to mitigate those effect, automated segmentation of the entire cortex and medulla could provide more representative and robust measurements.^27^ Finally, further developments might include semi-automated segmentation, an extension to T_2_* mapping^28^ to more sensitively assess oxygenation, and diffusion modules to further characterize fibrosis.

## Conclusion

We demonstrated that the proposed navigator-gated 2D radial GRE sequence PARMANav enables accurate and precise simultaneous T₁-T₂ mapping of the kidneys during free breathing at 3T. The navigator acceptance window width (NAWW) had minimal impact on the accuracy, although very narrow or wide windows introduced more outliers, which led to map degradation, potentialy due to residual motion and registration errors. Based on these findings, an NAWW of ±8 mm was selected as a trade-off between scan efficiency and motion robustness.

## Data Availability

All data produced in the present study are available upon reasonable request to the authors and agreement of the local ethics committee.

## Acknowledgement

This study was funded by the Swiss National Science Foundation (SNSF) under grant number CRSII5_202276.

